# The relationship between payer type and quality of care for women undergoing emergency cesarean section at three hospitals in rural Uganda

**DOI:** 10.1101/2024.03.17.24304434

**Authors:** Jonathan Mwiindi, Rigoberto Delgado, Lee Revere, Beth Wangigi, Edward Muguthu, Priscilla Busingye

## Abstract

**Background:** The study examined the relationship between payer type, and quality of care among mothers who deliver through emergency cesarean section in rural Ugandan hospitals.

**Methods:** We analyzed retrospective, de-identified patient data from three rural private-not-for-profit hospitals in Uganda. Two groups were included in the study, a self-payer patient group and a group fully sponsored by an international funding organization. The data was analyzed using hierarchical linear regression models comparing length of stay against payer type, and controlling for patient age, education level, parity, and indication for C-section. Length of stay (LOS) was assumed to represent a realistic proxy variable for patient quality of care.

**Results:** The self-pay group had statistically significant longer postoperative LOS (surgery to clinical discharge), and longer aggregate LOS, or admission to clinical discharge, compared to the sponsored group. Payer type was not significant in the admission-to-decision LOS, but payer type was highly significant for aggregate LOS (*p <* .001).

**Conclusion:** Case management in rural Ugandan hospitals influences quality of post-operative care for patients undergoing emergency C-sections. Expanding surgical funding, combined with effective case management approaches, is likely to increase quality of surgical care as measured by length of stay.

**Synopsis**: Post-operative quality of care for emergency C-section cases, measured by length of stay in Ugandan rural hospitals, is impacted by case management.

## Introduction

Deliveries by cesarean section (C-section) continue to rise globally accounting for 21% of all live childbirths; compared to only 7% in 1990 [1]. C-sections are essential in situations such as prolonged or obstructed labor, fetal distress, or abnormal presentation of the baby. However, access to these life-saving procedures is greatly determined by a patient’s geographical location [1]; the more rural a patient is located the harder it is to access surgical services [2]. About 8% of women in least developed countries give birth by caesarean section, and the rate decreases to 5% in sub-Saharan Africa [1]. Although in Uganda 88% of all the surgeries performed are related to pregnancy [3], and only 1 in 3 women who need a C-section receive one [4]; in rural areas such disproportion is estimated to be even greater [5].

Factors associated with lack of access to C-sections in most African countries, especially in rural settings, include faulty equipment, limited skilled healthcare workforce, unavailability of transportation (including a lack of money to pay for transport), inadequate health facilities, and delays in seeking emergency care [6] [7]. Another important factor is the perception of the patient population towards cesarean sections. A study by Nakimuli et al. (2015) concluded that increased fear of delivery via C-section among Ugandan women created delays in seeking required obstetric care [8]. Lastly, is the relative costs of these interventions which many families are unable to afford. Such factors, combined with delays at healthcare facilities, complicate conditions for many mothers resulting in the need for emergency C-section (ECS) [9];[10].

In Uganda, 62.5% of private and missionary hospitals provide the three most common surgeries in this country: C-section, laparotomy, and open fracture repair [2]. Improving access to healthcare services for low-income mothers in Uganda’s rural areas also depends on the involvement of different foundation programs, which offer support in the form of medical supplies, specialist expertise, or service-specific funding. Given that these support programs can play an important role in the health systems of the region, it is important to understand their impact in terms of health outcomes; particularly, there is limited research into the effect of programs which provide full cost coverage for ECS in low-income countries. The objective of this study was to examine the relationship between payer type (self-paying patients and patients whose care is supported) and quality of care among mothers who deliver through emergency cesarean section in rural Ugandan hospitals. The study uses the case of African Mission Healthcare (AMH), a foundation which partners with private-not-for-profit (PNFP) hospitals to cover costs of surgical care for indigent patients through its Surgery Access for Everyone program (SAFE). It is expected that the results of this study will serve to improve healthcare policy making to support rural hospitals.

## Methods

The study analyzed retrospective, de-identified patient data from three rural PNFP hospitals operating under the Uganda Catholic Medical Bureau (UCMB), Uganda. Two of these hospitals, Nyakibale and Rushoroza, formed part of the SAFE program, while the third, Ibanda Hospital, did not receive any SAFE benefits and was used as a control. The three hospitals are located in Southwestern Uganda, between 308 and 410 km from Kampala, the capital city. The three hospitals serve approximately 40,000 patients annually. Nyakibale is a 200-bed hospital with approximately 90 healthcare workers, Rushoroza is a 93-bed hospital with approximately 34 health workers, while Ibanda is a 178-bed hospital with 88 healthcare workers. The study data included two patient groups: 1) study group composed of patients whose delivery costs were covered entirely by the SAFE program given their extreme poverty conditions; and 2) the control, or self-payer group, composed of patients who had no health insurance and had all out-of-pocket expenses related to the delivery. In addition to covering the cost of ECS care, the SAFE program requires participating hospitals to comply with stipulated quality of care indicators, and accuracy of patient data collected. Quality checks are performed daily by personnel funded by SAFE, and comprise assessments of patient pre-operation profiles, post-operation feedback, and quarterly clinical audits. The patient sample was of convenience and included all records of women who had received emergency C-section in the three study hospitals between January and August 2020.

The statistical analysis consisted of frequency tables, mean differences, and used two hierarchical linear regression models assessing patient length of stay (LOS) against payer-type (self-pay and SAFE), and controlling for patient age, education level, parity (number of previous deliveries), and indication for C-section (fetal distress, previous scar, malpresentation and maternal compromise). Number of living children was not included in the models since this variable was not reported in the patient records reviewed. The dependent variable in the first regression model was decision to delivery interval (DDI), referring to the time between when the provider determines the patient requires a C-section and the time when the baby is delivered. The second model used as dependent a variable denominated as aggregate LOS, or the time lapse between a patient’s admission and clinical discharge. The term clinical discharge is used here to denote the event when a patient is considered able to leave the hospital only in terms of their health condition. Since hospitals in Uganda traditionally do not discharge a patient until full payment is made, it is important to distinguish between clinical discharge and actual hospital discharge. Thus, considering clinical discharge allowed us to estimate differences in post-operative LOS, controlling for differences in capacity to pay between the two patient groups. The purpose of modeling aggregate LOS was to evaluate separately the two elements that compose this variable: 1) the DDI; and 2) the postoperative (post-surgical) LOS. This breakdown facilitated the examination of quality service in terms of speed of service delivery (DDI), and to obtain metrics which can be used to benchmark against country averages of clinical and non-clinical aspects of post-surgical care (postoperative LOS). When surgical patients prolonged LOS, the risk of surgical site infections (SSI) increases [11].

Thus, LOS is a good proxy for measuring quality of surgical care. The shorter the LOS the better the quality of the service offered. In both regression models, LOS was assumed to represent a realistic proxy variable for quality of care [12]. DDI, for example, is an important risk factor of maternal mortality and poor neonatal outcomes [13]. All variables in the models were assessed for normality and multi-collinearity. The study used SPSS 2020 version 7.0 (SPSS Inc., Chicago, Ill., USA), and statistical significance was evaluated at the generally accepted level, α = .05.

Having Ibanda hospital, a non-SAFE institution, in the sample allowed to control for possible biases resulting from the quality checks performed by the SAFE program team members. The study received IRB approval from the University of Texas Health Science Center at Houston, Texas, USA, and from the Mbarara University of Science and Technology, Mbarara, Uganda. Given that the study involved retrospective, de-identified patient data, informed consent was waived.

## RESULTS

As shown in table 1, a total of patient 771 records were included in this study; thirty five (35) were excluded from the analysis due to incomplete data leaving 736 records. Of these, 378 were in the Self-payer group and 358 were part of the SAFE ECS program. Ibanda hospital only had patients from the Self-pay group (291 patients), while Rushoroza and Nyakibale hospitals had both patient groups represented. Majority of the participants were married (Self-pay 68%; SAFE, 95%), and mostly received an emergency C-section (Self-pay 91%; SAFE, 98%). The mean average age was 26 (SD 6.17) and 27 (SD 6.35) for the Self-pay and SAFE groups, respectively. Maternal compromise (Self-pay 30%, SAFE 22%) and previous scar (Self-pay 28%, SAFE 33%) were the most common indications for emergency C-sections. In addition, there were no fetal losses for the patients in this cohort and nearly all (99%) did not have a surgical site infection.

**Table 1:** General Characteristics of the Study Participants, by Payer Group.

| Characteristic | Self-Pay<br>*n* = 378 | SAFE<br>*n* = 358 |
| --- | --- | --- |
| Study site |  |  |
| Ibanda | 291 (77%) | 0 (0%) |
| Nyakibale | 69 (18%) | 219 (61%) |
| Rushoroza | 18 (5%) | 139 (39%) |
| Marital status |  |  |
| Married | 256 (68%) | 341 (95%) |
| Single | 0 (0%) | 12 (3%) |
| Separated | 0 (0%) | 5 (1%) |
| Missing | 122 (32%) | 0 (0%) |
| Surgery Type |  |  |
| Emergency C-section | 344 (91%) | 350 (98%) |
| Elective C-section | 33 (9%) | 8 (2%) |
| Indication of C-section |  |  |
| Maternal Compromise | 112 (30%) | 77 (22%) |
| Previous Scar | 104 (28%) | 118 (33%) |
| Dystocia | 66 (17%) | 63 (18%) |
| Malpresentation | 51 (13%) | 55 (15%) |
| Fetal Distress | 45 (12%) | 45 (13%) |
| Surgical Site Infection |  |  |
| No | 376 (99%) | 355 (99%) |
| Yes | 2 (1%) | 3 (1%) |
| Birth outcome |  |  |
| Alive | 355 (94%) | 343 (96%) |
| Dead | 23 (6%) | 13 (4%) |

*Note.* Due to rounding errors, column-wise percentages may not equal 100%.

The results from mean differences analysis, presented in Table 2, indicates shows that there were statistically significant differences between payer types for postoperative LOS, or surgery to discharge time (*p* < .001), and aggregate LOS; the time between admission and clinical discharge (*p* < .001). This indicates that the Self-pay group on average had significantly longer postoperative and aggregate LOS in comparison to the SAFE payer group, but the same mean DDI compared to SAFE patients. There was no statistical difference in mean age, number of previous deliveries, or DDI between the two groups.

**Table 2:** Analysis of Payer Groups Mean Differences for Selected Patient Characteristics.

| Variable | *x* | *SD* | *n* | Min | Max | *t*(734) | *p* |
| --- | --- | --- | --- | --- | --- | --- | --- |
| Patient Age |  |  |  |  |  | -1.71 | .088 |
| Self-Pay | 26.32 | 6.17 | 378 | 13.00 | 47.00 |  |  |
| SAFE | 27.11 | 6.35 | 358 | 15.00 | 43.00 |  |  |
| Number of previous deliveries |  |  |  |  |  | 0.61 | .542 |
| Self-Pay | 2.80 | 1.91 | 378 | 1.00 | 13.00 |  |  |
| SAFE | 2.71 | 1.82 | 358 | 1.00 | 9.00 |  |  |
| DDI - admission to surgery (hours) |  |  |  |  |  | 1.05 | .294 |
| Self-Pay | 16.47 | 23.53 | 378 | 0.12 | 136.33 |  |  |
| SAFE | 14.70 | 22.07 | 358 | 0.00 | 117.45 |  |  |
| Postoperative LOS – surgery to discharge (hours) |  |  |  |  |  | 16.77 | <.001 |
| Self-Pay | 98.89 | 27.48 | 378 | 18.08 | 251.80 |  |  |
| SAFE | 71.24 | 15.17 | 358 | 9.78 | 154.25 |  |  |
| Aggregate LOS – admission to discharge (hours) |  |  |  |  |  | 12.82 | <.001 |
| Self-Pay | 115.36 | 34.34 | 378 | 33.50 | 255.83 |  |  |
| SAFE | 85.94 | 27.30 | 358 | 12.00 | 250.33 |  |  |

*Note.* *x* = mean; *SD* = standard deviation, *n* = sample size, *t* = statistic from the independent samples t-test

### Decision-Delivery Interval - Admission to Surgery

Table 3 presents the results of the hierarchical linear regression analysis predicting DDI (admission to surgery) against payer type. The three models (steps 1-3) were statistically significant, and in all steps the signs of the estimated coefficients, and degree of statistical significance, remained the same for all variables supporting the observations presented in this section. The explained variation (*R*^2^) was relatively small in all three models. In Step 1, *F* (2, 733) = 3.65, *p*=0.026 (*R*^2^ = 0.010), patient age showed significant statistical evidence of association with DDI (*β* = 0.11, *p* = .029), indicating that older patients reported longer hours between admission and delivery compared to younger patients. The number of previous deliveries showed a highly significant relationship with DDI (*β* = −0.14, *p* = .008), suggesting that number of previous deliveries was inversely related to number of hours a patient experienced on the DDI.

**Table 3:**
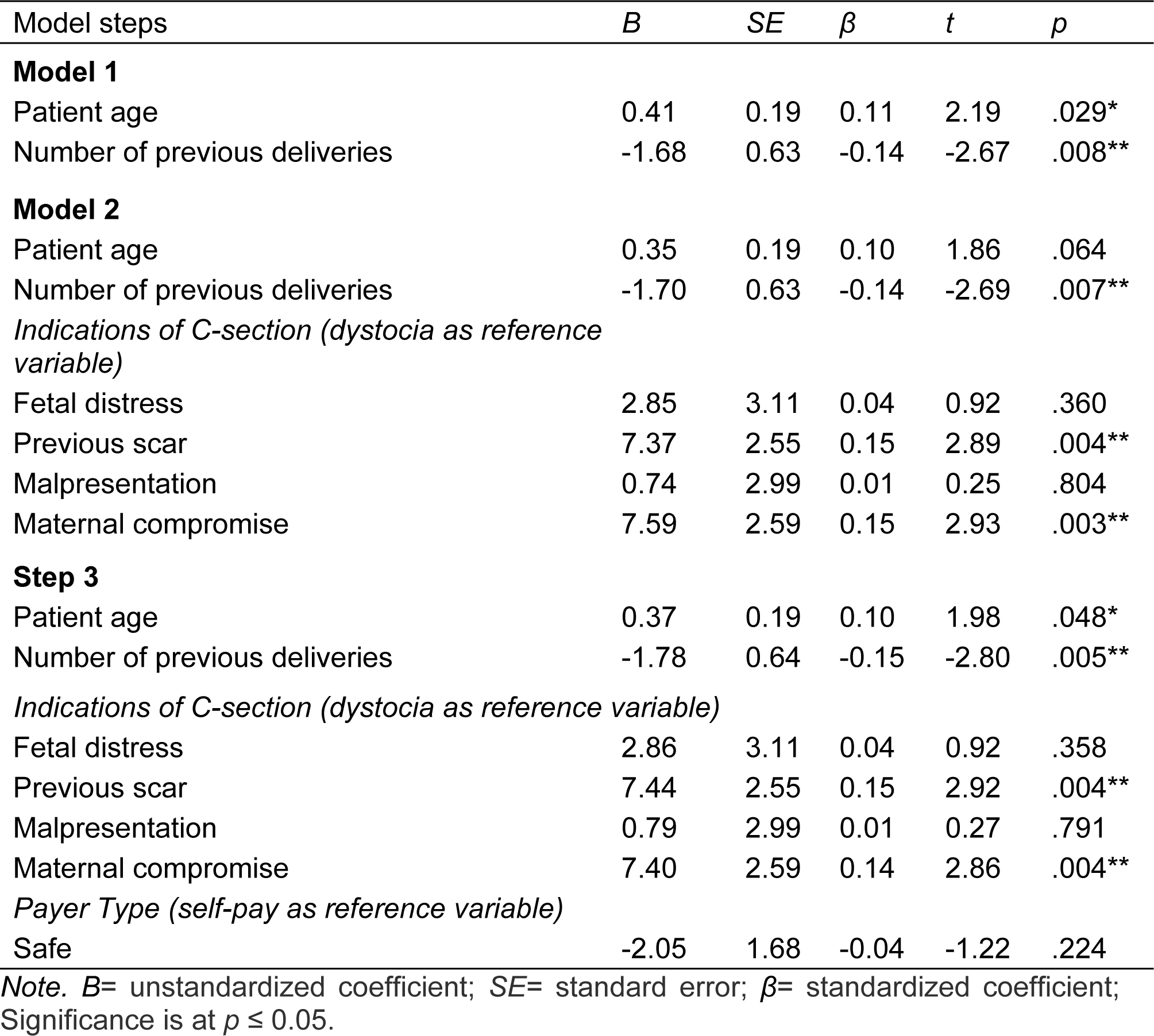
Hierarchical Regression Analysis Predicting DDI.

| Model steps | *B* | *SE* | $\beta$ | *t* | *p* |
| --- | --- | --- | --- | --- | --- |
| **Model 1** |  |  |  |  |  |
| Patient age | 0.41 | 0.19 | 0.11 | 2.19 | .029* |
| Number of previous deliveries | -1.68 | 0.63 | -0.14 | -2.67 | .008** |
| **Model 2** |  |  |  |  |  |
| Patient age | 0.35 | 0.19 | 0.10 | 1.86 | .064 |
| Number of previous deliveries | -1.70 | 0.63 | -0.14 | -2.69 | .007** |
| *Indications of C-section (dystocia as reference variable)* |  |  |  |  |  |
| Fetal distress | 2.85 | 3.11 | 0.04 | 0.92 | .360 |
| Previous scar | 7.37 | 2.55 | 0.15 | 2.89 | .004** |
| Malpresentation | 0.74 | 2.99 | 0.01 | 0.25 | .804 |
| Maternal compromise | 7.59 | 2.59 | 0.15 | 2.93 | .003** |
| **Step 3** |  |  |  |  |  |
| Patient age | 0.37 | 0.19 | 0.10 | 1.98 | .048* |
| Number of previous deliveries | -1.78 | 0.64 | -0.15 | -2.80 | .005** |
| *Indications of C-section (dystocia as reference variable)* |  |  |  |  |  |
| Fetal distress | 2.86 | 3.11 | 0.04 | 0.92 | .358 |
| Previous scar | 7.44 | 2.55 | 0.15 | 2.92 | .004** |
| Malpresentation | 0.79 | 2.99 | 0.01 | 0.27 | .791 |
| Maternal compromise | 7.40 | 2.59 | 0.14 | 2.86 | .004** |
| *Payer Type (self-pay as reference variable)* |  |  |  |  |  |
| Safe | -2.05 | 1.68 | -0.04 | -1.22 | .224 |

*Note.* *B*= unstandardized coefficient; *SE*= standard error; $\beta$ = standardized coefficient; Significance is at $p \leq 0.05$ .

In Step 2, *F* (6, 729) = 3.80, *p* = 0.001, (*R*^2^ =.030), the number of previous deliveries remained highly significant (*β* = −0.14, *p* = .007). In addition, two indications for C-section were highly significant; previous scars (*β* = 0.15, *p* = .004) and maternal compromise (*β* = 0.15, *p* = .003), suggesting that these patients had longer DDI compared to those showing only conditions of dystocia, the reference variable. In Step 3, *F* (7, 728) =3.47, *p* =0.001, (*R*^2^ =0.032), patient age showed statistical significance (*β* = −0.10, *p* = .048), while number of previous deliveries (*β* = −0.15, *p* = .005), previous scars (*β*= 0.15, *p* = .004), and maternal compromise (*β*= 0.14, *p* = .004) were highly significant as factors of DDI variation. Fetal distress, and malpresentation showed no evidence of statistical relationship with DDI, and as compared to the reference variable dystocia. Further, no significant difference was found between women in the SAFE group compared to the Self-pay group regarding DDI.

### Aggregate LOS - Admission to Discharge

Table 4 presents the results of the hierarchical linear regression analysis predicting aggregate LOS against payer type. The model in Step 1 was not significant with none of its two variables indicating statistical significance. In Step 2, *F* (6, 729) = 2.42, *p* = .025, (*R*^2^ = .020), maternal compromise (*β*= 0.15, *p* = .003) showed highly significant association with aggregate LOS compared to dystocia. This finding indicates that patients with maternal compromise remained for a longer period between admission and clinical discharge compared to those patients without maternal compromise.

**Table 4:** Hierarchical Regression Analysis Model Predicting Aggregate LOS.

| Model steps | *B* | *SE* | $\beta$ | *t* | *p* |
| --- | --- | --- | --- | --- | --- |
| **Model 1** |  |  |  |  |  |
| Patient age | -0.26 | 0.28 | -0.05 | -0.93 | .355 |
| Number of previous deliveries | 0.26 | 0.96 | 0.01 | 0.27 | .785 |
| **Model 2** |  |  |  |  |  |
| Patient age | -0.29 | 0.28 | -0.05 | -1.02 | .309 |
| Number of previous deliveries | 0.50 | 0.96 | 0.03 | 0.52 | .604 |
| *Indications of C-section (dystocia as reference variable)* |  |  |  |  |  |
| Fetal distress | 4.91 | 4.71 | 0.05 | 1.04 | .298 |
| Previous scar | 3.46 | 3.86 | 0.05 | 0.90 | .370 |
| Malpresentation | -1.10 | 4.53 | -0.01 | -0.24 | .808 |
| Maternal compromise | 11.60 | 3.92 | 0.15 | 2.96 | .003** |
| **Model 3** |  |  |  |  |  |
| Patient age | 0.07 | 0.26 | 0.01 | 0.26 | .796 |
| Number of previous deliveries | -0.61 | 0.87 | -0.03 | -0.70 | .485 |
| *Indications of C-section (dystocia as reference variable)* |  |  |  |  |  |
| Fetal distress | 5.08 | 4.28 | 0.05 | 1.19 | .235 |
| Previous scar | 4.48 | 3.50 | 0.06 | 1.28 | .201 |
| Malpresentation | -0.37 | 4.11 | -0.00 | -0.09 | .929 |
| Maternal compromise | 8.96 | 3.56 | 0.11 | 2.52 | .012* |
| *Payer Type (self-pay as reference variable)* |  |  |  |  |  |
| Safe | -29.07 | 2.31 | -0.42 | -12.56 | < .001** |

*Note.* *B*= unstandardized coefficient; *SE*= standard error; $\beta$ = standardized coefficient; Significance is at $p \leq 0.05$ .

In Step 3, *F* (7, 728) = 25.04, *p* < .001, (*R*^2^ = .194), both maternal compromise (*β*= 0.11, *p* = .012) and payer type (*β*= −0.42, *p <* .001) showed significant and highly significant association, respectively, with aggregate LOS. This finding provides strong evidence that patients under the SAFE program spent considerably less time in hospital compared to those patients in the Self-pay group. In both models the signs of the coefficient and degree of significance for all variables remained the same. The coefficient of determination (*R*^2^) in the last model was considerably higher accounting for almost 20% of variation.

## Discussion

This study investigated the relationship between payer type and the length of stay (LOS) of mothers who deliver by emergency C-section in rural hospitals of Uganda. Two patient groups formed part of the study: the Self-pay, patients paying entirely out-of-pocket for medical services; and the group supported by the Surgery Access for Everyone (SAFE) program managed by the foundation African Mission Healthcare. On average, patients in the SAFE group had shorter aggregate LOS than patients in the Self-pay group, considering both time intervals of surgery to clinical discharge (71.24 vs 98.89 hrs. for SAFE and Self-pay patients, respectively), and aggregate LOS, or admission to clinical discharge (85.94 hrs. vs. 115.36 hrs.). Thus, a woman paying for herself was more likely to spend an additional 29 hours in the hospital than a woman whose emergency C-section was sponsored.. Since the mean decision-to delivery time interval (DDI) was statistically the same between the Self-pay and SAFE patient groups, differences in aggregate LOS between the two payer types are likely due to mean length differences in the post-discharge LOS (surgery to clinical discharge). This statement is also supported by the results from the regression analysis. Factors such as patient age, previous deliveries, and indications of C-section played a significant role explaining DDI variation, however, payer type was not a significant variable (Table 3). In contrast, payer type was a significant variable explaining variation in aggregate LOS, along with maternal compromise (Table 4).

Given that DDI provides a broad indicator of performance during a critical level of surgical care, statistically similar estimates of mean DDI likely indicate that patients in both payer groups receive similar quality treatment during a C-section. Post-surgical care, on average, involves less urgent medical attention, and mean statistical differences in this metric could reflect dissimilarities in quality of recovery care and case management. One important feature of the SAFE program, which helps explain lower LOS for its beneficiaries, is its process of an intensive approach to case management. The case management is carried out by a hospital employee. Case management involves communicating with clinicians to help ensure a patient’s recovery progress, handling complaints, and guiding patients through the discharge process. While the case manager is not likely to be involved in the DDI process, the systematic case management during the post-surgery period likely contributes to reducing aggregate LOS.

The observed average DDI mean estimates for both Self-pay and SAFE patient groups (16.47 hrs. and 14.70 hrs., respectively) are consistent with findings from other studies in Uganda - DDI ranges of 12:00 - 20:00 hours [14] - and neighboring Tanzania and Kenya - only 12% and 3% of hospitals in these two countries, respectively, had DDI time frames as low as 75-minutes [13][15]. These findings, however, underline an ongoing challenge regarding C-section procedures in Uganda, if compared to the 30-min DDI benchmark recommended in developed countries [16]. DDI differences may be due to general infrastructure and economic factors such as a lack of surgical funds and a skilled health worker to patient ratio [17], or severe lack of medication, equipment, and operating rooms [18]. These factors highlight the fact that there are no DDI recommendations for low- and middle-income countries like Uganda [19–21], and the results of this study are contributing to quantifying current DDI time frames.

The study encountered some limitations. First, due to data constraints, estimates of DDI were calculated considering time between admission and surgery, and not from when the decision for a C-section and to when the surgery took place. This could have resulted in an overestimation of DDI for both payer groups. Second, the clinical discharge time was not indicated in the patient charts in Ibanda and Rushoroza hospitals. Instead, clinical discharge time for patients in these hospitals was considered to be 9:00am, the start of general rounds. This may have underestimated post-surgery LOS in the control arm. There are opportunities for further research, including measuring long-term patient quality of life differences between payer types. Also, there is the need to study the association between LOS and the referring facility post-discharge to measure effectiveness of patient follow-up.

## Conclusion

The findings of this study highlight a significant difference in LOS between mothers whose ECS are sponsored by payers compared to mothers who pay out-of-pocket -- using clinical discharge time to control for ability to pay. No differences in surgical outcomes were found between payer groups, but a proactive case management approach was likely the key factor contributing to differences in post-surgery quality of care. Therefore, funding for ECS should be expanded, especially for poor, rural women who would otherwise have limited access surgery.

## Data Availability

All relevant data are within the manuscript and its supporting information files

## Acknowledgments

The authors thank Dr. Joseph Kolars, Dr. Wendy Nyamusana, Sr. Gertrude Kabanyomozi, Sr. Miria Mazire, and Sr. Mary Mangdalene for their assistance in the collection of data and revisions of the manuscript.

## Author Contributions

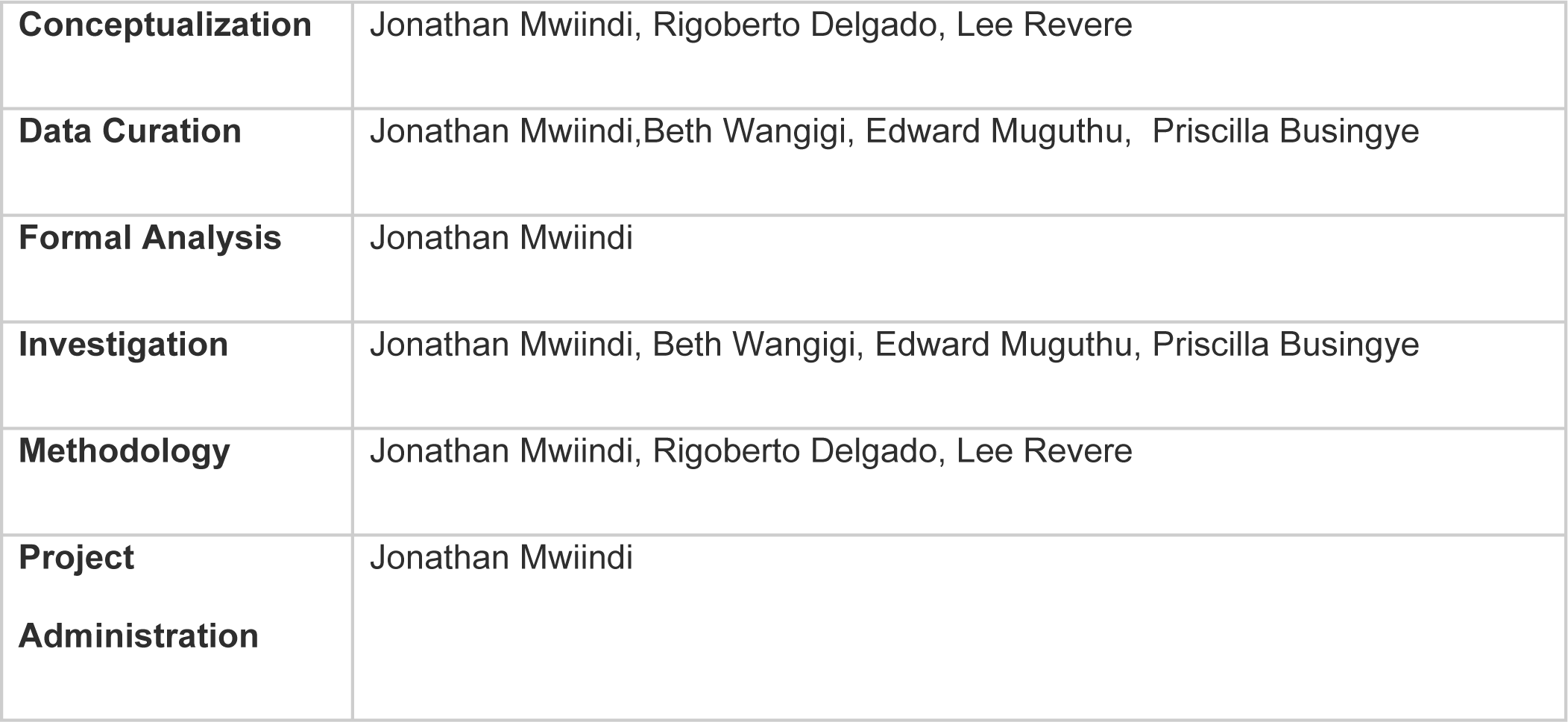
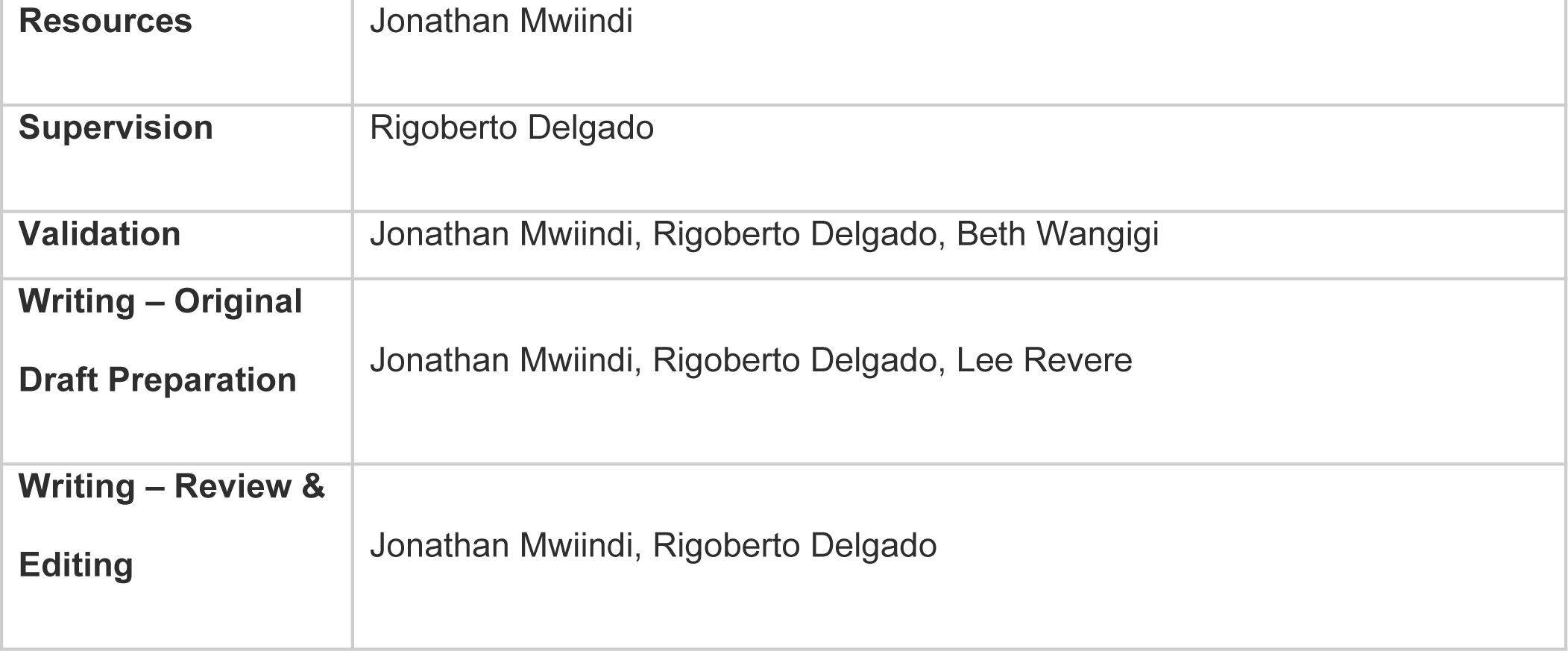

